# Stress, Burnout and Depression in Women in Healthcare during COVID-19 Pandemic: Rapid Scoping Review

**DOI:** 10.1101/2020.07.13.20151183

**Authors:** Abi Sriharan, Savithiri Ratnapalan, Andrea C. Tricco, Doina Lupea, Ana Patricia Ayala, Hilary Pang, Dongjoo Daniel Lee

**Affiliations:** Institute of Health Policy, Management and Evaluation, Dalla Lana School of Public Health, University of Toronto; Department of Pediatrics and Dalla Lana School of Public Health, University of Toronto. Hospital for Sick Children, Toronto; Li Ka Shing Knowledge Institute, St. Michael’s Hospital, Unity Health Toronto; Epidemiology Division and Institute of Health Policy, Management and Evaluation, Dalla Lana School of Public Health, University of Toronto, Queen’s Collaboration for Health Care Quality Joanna Briggs Institute Centre of Excellence, Queen’s University; Ontario Medical Association, Physician Health Program; Gerstein Science Information Centre, University of Toronto; Faculty of Medicine, Institute of Health Policy, Management, and Evaluation, University of Toronto

**Keywords:** Coronavirus, COVID-19, women in health care, stress, burnout, depression

## Abstract

**Objectives:** The overall objectives of this rapid scoping review are to (a) synthesize the common triggers of stress, burnout, and depression faced by women in health care during the COVID-19 pandemic, and (b) identify individual-, organizational-, and systems-level interventions that can support the well-being of women HCWs during a pandemic.

**Design:** This scoping review is registered on Open Science Framework (OSF) and was guided by the JBI guide to scoping reviews and reported using the Preferred Reporting Items for Systematic reviews and Meta-Analysis (PRISMA) extension to scoping reviews. A systematic search of literature databases (Medline, EMBASE, CINAHL, PsycInfo and ERIC) was conducted from 2003 until June 12, 2020. Two reviewers independently assessed full-text articles according to predefined criteria.

**Interventions:** We included review articles and primary studies that reported on stress, burnout, and depression in HCWs; that primarily focused on women; and that included the percentage or number of women included. All English language studies from any geographical setting where COVID-19 has affected the population were reviewed.

**Primary and secondary outcome measures:** Studies reporting on mental health outcomes (e.g., stress, burnout, and depression in HCWs), interventions to support mental health well-being were included.

**Results:** Of the 2,803 papers found, 31 were included. The triggers of stress, burnout and depression are grouped under individual-, organizational-, and systems-level factors. There is a limited amount of evidence on effective interventions that prevents anxiety, stress, burnout and depression during a pandemic.

**Conclusions:** Our preliminary findings show that women HCWs are at increased risk for stress, burnout, and depression during the COVID-19 pandemic. These negative outcomes are triggered by individual level factors such as lack of social support; family status; organizational factors such as access to personal protective equipment or high workload; and systems-level factors such as prevalence of COVID-19, rapidly changing public health guidelines, and a lack of recognition at work.

**Strengths and limitations of this study:**

- A rapid scoping review was conducted to identify stress, burnout and depression faced by women HCWs during COVID-19.
- To ensure the relevance of our review, representatives from the women HCWs were engaged in defining the review scope, developing review questions, approving the protocol and literature search strategies, and identifying key messages.
- It provides a descriptive synthesis of current evidence on interventions to prevent mental health for women HCWs.
- Most studies used cross-sectional surveys, making it difficult to determine the longitudinal impact.
- There was significant variability in the tools used to measure mental health.

## INTRODUCTION

COVID-19 pandemic-related measures, such as prolonged periods of social isolation, unexpected employment disruptions, school closures, financial distress, and changes to routine, are having an unprecedented negative impact on women’s mental well-being (UN). Over 80% of the health workers in Canada are women (Porter et al, 2017). Women in health care already face systemic challenges related to workplace gender biases, discrimination, sexual harassment, and other inequities (Ghebreyesus, T, 2019). Studies show that women physicians are more likely than male physicians to experience depression, burnout, and suicidal ideation (Gold, et al, 2016, Guille, C et al). Additionally, women perform three times more unpaid care work than men as parents and primary caregivers to family members (UN).

The COVID-19 pandemic has led to increased psychological trauma and suicide among health care workers (HCWs) (Mock, J, 2020., Orr, C., 2020). A poll of HCWs conducted by the Public Health Agency of Canada in April 2020 showed that 47% of respondents expressed the need for psychological support due to COVID-19 related factors; 90% of the respondents were women (McKinley, S. 2020). Similarly, a survey conducted by the British Medical Association in April, 2020 of HCWs showed that 44% of respondents indicated they were experiencing burnout, depression, anxiety, or other mental health conditions due to COVID-19-related factors (BMA, 2020). Unaddressed stress and burnout can lead to depression, suicidal ideation, and substance abuse (Oreskovich, et al 2012, Guille et al, 2017). A healthy workforce is the cornerstone of a well-functioning health care system. Yet, there is a systemic lack of evidence-informed services that provide timely, accessible, and high-quality care for HCWs during public health crises. This is especially relevant for health systems and professional societies who recognize the importance of preventing and mitigating stress, burnout, depression, and suicidal ideation in their workforce during pandemics. In addition, these interventions are essential for the wellbeing and retention of the health care workforce. This review attempts to answer the following questions: What are the common triggers of stress, burnout, and depression faced by women in health care during the COVID-19 pandemic? What individual-, organizational-, and systems-level interventions can support the well-being of women HCWs during a pandemic?

### Overall Objectives

The overall objectives of this review are to (a) synthesize the common triggers of stress, burnout, and depression faced by women in health care during the COVID-19 pandemic, and (b) identify individual-, organizational-, and systems-level interventions that can support the well-being of women HCWs during a pandemic.

## METHODS

### Commissioning agency

The Canadian Institute for Health Research issued a special call to address COVID-19 in Mental Health & Substance Use issues. In order to provide decision-makers with timely results, a rapid scoping review was conducted in accordance with the WHO Rapid Review Guide and the JBI 2020 guide to scoping reviews (Peters, MDJ, 2020, Tricco, A et al 2017) and reported using Preferred Reporting Items for Systematic Reviews and Meta-Analyses (PRISMA) for scoping reviews.

### Protocol

A protocol for this review was developed in collaboration with the women HCW (SR), primary knowledge user (DL), rapid review methodologist (AT) and a leadership wellness coach (AS). This review is registered with the Open Science Framework (https://osf.io/y8fdh/?view_only=1d943ec3ddbd4f5c8f6a9290eca2ece7).

### Eligibility Criteria

The following PICOS (population, intervention, comparator, outcome, Study Design) eligibility criteria were developed a priori:

#### Population

Women HCWs. We define HCWs as “all people engaged in actions whose primary intent is to enhance health,” (WHO, 2006). This encompasses a broad array of health workers, including doctors, nurses, pharmacists, midwives, paramedics, physical therapists, technicians, personnel support workers, and community health workers. We included studies that primarily focused on women.

#### Interventions

Our inclusion criteria were all studies (primary and review articles) that reported on the causes of stress, burnout, and depression in HCWs and/or reported programs to mitigate stress, burnout, and. Depression in HCWs.

#### Comparators

Not Applicable for the purpose of this scoping review.

#### Outcomes

We looked at following outcomes: stress, burnout, and depression. We define stress as the degree to which one feels overwhelmed and unable to cope as a result of unmanageable pressures (Maslach, 1962). We define burnout as the experience of emotional exhaustion, depersonalization, or cynicism, along with feelings of diminished personal efficacy or accomplishment in the context of the work environment (Templeton, 2019). We characterize depression according to a series of symptoms, including low mood, changes in appetite and sleep, difficulty concentrating, loss of interest/pleasure and thoughts of suicide that persist for at least two weeks (Mental Health Foundation).

#### Study Design

We included review articles and primary studies where data were collected and analyzed using quantitative, qualitative, and mixed methods. We excluded editorials and opinion pieces unless the authors shared their personal experiences.

### Search methods and information sources

We conducted comprehensive search strategies in the following electronic databases: Medline (via OVID), Embase (via Ovid), CINAHL (via EBSCOHost), PsycINFO (via Ovid), and ERIC (via ProQUEST). Search strategies were developed by an academic health sciences librarian (APA), with input from the research team. The search was original built in MEDLINE Ovid, and peer reviewed using the Peer Review of Electronic Search Strategies (PRESS) tool (Mcgowan et al 2016), before being translated into other databases using their command language if applicable. The Coronavirus (Covid-19) 2019-nCov expert search from Ovid MEDLINE was used and translated to other databases. Searches were limited by date from 2003-June 12^th^, 2020, and by English language. The final search results were exported into Covidence, a review management software, where duplicates were identified and removed.

### Screening Process

To minimize selection bias, we piloted 20 articles against *a priori* inclusion and exclusion criteria. Each article title was reviewed by two independent screeners against using Covidence. A third reviewer reviewed conflicts and resolved disagreements through discussion. Two reviewers also independently screened the full text of potentially eligible articles to check whether the articles fulfilled the inclusion criteria.

### Data Charting

We used a predefined data extraction form to extract data from the papers included in the review. To ensure the integrity of the assessment, we piloted the data extraction form on three studies. We extracted the following information from the studies: the first author, year of publication, HCWs enrolled in the study, geographic location, study methods, and intervention information that could help answer our objectives. We did not appraise quality or risk of bias of the included articles, consistent with accepted scoping review methods (Peters MDJ et al, 2020 and Tricco, A et al 2017). Ethical approval was not required for this review.

### Data Synthesis

Due to heterogeneity regarding outcome measurement and statistical analysis, data was descriptively synthesized.

### Patient involvement

No patients were involved in setting the research question or the outcome measures, nor were they involved in the design and implementation of the study.

## RESULTS

### Search Results

The search resulted in a total of 3,633 records. After 830 duplicates were removed, 2,803 records remained to be screened. We excluded 2,279 records based on title and abstract screening. We assessed 524 full-text articles. Most of these articles are opinion pieces and commentaries. Thirty-one published studies met our inclusion criteria and were included in this review.

### Characteristics of Studies

Our search identified 31 eligible studies; 3 of these were review articles (Table 1), 26 of these studies focused on the prevalence of mental health issues in health care professionals (Table 2). Two studies were case studies (Table 2). Sixteen of the primary studies were conducted in China, whereas others were conducted in Saudi Arabia, Italy, Singapore, India, and Colombia. These studies primarily focused on doctors, nurses, and generalized groups of allied health professionals. One study focused on dentists, whereas another focused-on pharmacists. The study samples included both male and women health professionals. Only one study focused exclusively on women in health care (Li, G, 2020). Anxiety, depression, stress/distress symptoms, post-traumatic stress disorder, and insomnia were commonly assessed mental health issues in these studies.

**Table 1:** Summary of Evidence from the Systematic Reviews

| Author | Study Type | Review Focus | No of HCWs | No of studies included | Dates Covered | Key findings |
| --- | --- | --- | --- | --- | --- | --- |
| Pappa et al. 2020 | Systematic review and meta-analysis | Prevalence of depression, anxiety, and insomnia in health care workers (HCWs) during the COVID-19 pandemic | 33,062 | 13 | Upto April 2020 | Increased level of insomnia, depression and anxiety observed in HCWs during COVID-19. |
| Spoorthy, et al 2020 | Review of literature | Review of mental health problems faced by HCWs during the COVID-19 pandemic | 3,043 | 6 | Upto April 2020 | COVID-19 increased stress level in HCWs. Gender, profession and social support are associated with higher rates of stress, anxiety, depressive symptoms, and insomnia in HCWs. |
| Vindegaard, et al , 2020 | Systematic Review | Review of measured psychiatric symptoms among infected patients compared to noninfected psychiatric patients, HCWs, and non-HCWs | 26,877 | 20 | Upto May 2020 | HCWs have increased depression or depressive symptoms, anxiety, psychological distress, and poor sleep quality. The women gender, poor-self-rated health, and relatives with COVID-19 are associated with low psychological well-being. |

**Table 2:** Summary of Primary Studies

| Author | Study Sample | Study Type | Female | Doctors | Nurses | Other HCWs | Country |
| --- | --- | --- | --- | --- | --- | --- | --- |
| 1. Al Sulais | 529 | Cross-sectional Survey (CSS) | Breakdown N.A | X | - | - | Saudi Arabia |
| 1. Almagharabi | 1036 | CSS |  |  |  |  | Saudi Arabia |
| 2. Cai | 534 | CSS | 367 | X |  |  | China |
| 3. Chew | 906 | CSS | Breakdown Not Available (N.A) | X | (N.A.) | (N.A.) | China |
| 4. Chowdhury, S,M |  | Qualitative |  |  |  |  |  |
| 5. DeStefani | 1500 |  | 836 | - | - | Dentists | Italy |
| 6. Elbay | 442 | CSS | 251 | X | - | - | Turkey |
| 7. Felice | 388 | CSS | 235 | X | - | X | Italy |
| 8. Huang | 600 | CSS | 305 | X | X | X | China |
| 9. Kang | 994 | CSS | 850 | X | X | - | China |
| 10. Karasneh | 486 | CSS | 382 | - | - | X | Jordan |
| 11. Khanna | 2355 | CSS | 1332 | X | - | - | India |
| 12. Lai | 1257 | CSS | 964 | X | X | - | China |
| 13. Li | 5317 | CSS | 5317 | X | X | X | China |
| 14. Liu | 512 | CSS | 433 | N.A. | N.A. | N.A. | China |
| 15. Marton |  | Qualitative |  |  |  |  |  |
| 16. Pedrozo-Papa | 179 | CSS | Breakdown N.A | X | X | X | Spain |
| 17. Romero | 1671 | CSS | Breakdown N.A | X | - | - | Spain |
| 18. Song | 14825 | CSS | 9536 | X | X | - | China |
| 19. Sun | 442 | CSS | 368 | X | X | X | China |
| 20. Uzun | 103 | CSS | 91 | X | X | X | Turkey |
| 21. Wu | 190 | CSS | 157 | - | X | X | China |
| 22. Xiao | 958 | CSS | 644 | X | X | X | China |
| 23. Yin | 371 | CSS | 228 | X | X | X | China |
| 24. Yuan | 939 | CSS | 582 | X | - | X | China |
| 25. Zhang | 304 | CSS | 178 | N.A. | N.A. | N.A. | China |
| 26. Zhang | 927 | CSS | 678 | X | X | X | China |
| 27. Zhu | 165 | CSS | 137 | X | X | - | China |

A variety of assessment tools were used to measure mental health in these studies. Common tools used to measure psychosocial well-being included DASS-21, Impact of Event Scale Revised Questionnaire (IES-R), Connor-Davidson Resilience Scale, Chinese Perceived Stress Scale, Patient Health Questionnaire-9 (PHQ-9), Generalized Anxiety Disorder (GAD-7) Scale, Questionnaire Star, Psychological Symptom Screening Test (SCL-90-R), Beck Anxiety Inventory and Short Psychiatric Rating Scale, Maslach Burnout Inventory-Medical Personnel, Perceived Stress Scale and Hospital Anxiety/Depression Scale, Post-traumatic Stress Disorder Checklist for DSM-5 and the Pittsburgh Sleep Quality Index, Stress Response Questionnaire, Zung Self-Rating Anxiety Scale SF-12, K6, Insomnia Severity Index, Self-Rating Depression Scale, and Simplified Coping Style Questionnaire.

### Common Triggers of Stress, Burnout, and Depression Faced by Women in Health Care During the Coronavirus Pandemic

Common triggers of mental health issues were fears of getting infected with COVID-19 and putting family members at risk (Al Sulais,et al, 2020, Almaghrabi, et al, 2020, Cai et al, 2020), as well as concerns about professional growth, difficulty meeting living expenses (Khanna et al, 2020), and having family members with suspected and confirmed COVID-19 (Li, et al, 2020). Individual-, organizational-, and systems-level factors are reported as common triggers of stress, burnout, and depression in women HCWs.

#### Individual-level factors

Women HCWs are more likely than men HCWs to experience psychological stress and burnout (Al Sulais et al, 2020, Huang, et al, 2020., Elbay,et al, 2020., Xiao, et al, 2020., Yin, et al, 2020., Yuan, et al 2020., Zhu, et al, 2020). More specifically, young women HCWs and mid-career women HCWs were more likely to experience emotional and mental health issues due to COVID-19 (Elbay,et al, 2020., Li, et al., 2020, Song et al., 2020). Similarly, less working experience and self-perception about lack of competency to care for COVID-19 patients was associated with increased prevalence of stress and burnout (Elbay, et al., 2020, Song et al., 2020). Women who are single or lacking social support are more at risk of developing symptoms of anxiety, stress and burnout (Song et al, 2020, Elbay,et al, 2020, Kang, et al, 2020, Li, et al., 2020, Uzun et al.,). Women HCWs with medical or psychiatric comorbidities (Li, et al, 2020, Uzun et al., 2020) or increased alcohol use are at higher risk of mental health issues (Song et al.2020). Surprisingly, women HCWs who have more than two children experience higher prevalence of psychosocial well-being (Elbay, et al, 2020).

#### Organizational-level factors

Long working hours and increased workload (Elbay,et al, 2020, Song, et al, 2020, Felice, et al, 2020); increased number of COVID-19 patients under their care (Elbay, et al, 2020, Liu, et al, 2020,); lack of access to personal protective equipment (Cai, et al, 2020, Felice, et al, 2020, Huang, et al, 2020, Xiao, et al, 2020, Zhang, et al, 2020, Almaghrabi, et al, 2020, De Stefani et al, 2020); lack of infection control guidelines and protocols (Elbay, et al, 2020, Cai, et al, 2020, De Stefani, et al, 2020, Wu et al, 2020); lack of support and recognition by their peers, supervisors, and hospital leadership (Elbay, et al, 2020, Cai et al, 2020); and work location (Zhang et al, 2020, Xiao et al, 2020) are reported as common triggers of mental health issues related to the work environment.

#### Systems-level factor

Increased incidence of COVID-19 cases in the local area (Cai, et al, 2020), changes in public health measures and guidelines (Pedrozo et al, 2020), information shared in the media (Karansnel et al, 2020), and lack of recognition by the government officials and policy makers of HCWs’ work conditions (Cai, 2000) are reported to increase stress and mental health issues among HCWs.

### Interventions That Can Support the Well-Being of Women HCWs During a Pandemic

Very few studies have discussed potential interventions to support women in health care with COVID-19 related stress, anxiety, and mental health. Women with increased workloads preferred to use psychological support (Felice et al, 2020). Regular exercise is considered a protective factor for depression and anxiety (Li et al, 2020). Time is considered a modifiable factor that improves anxiety level (Yuan et al, 2020). Mental health services such as online resources, psychological assistance hotlines, and group activities for stress reduction are poorly utilized by HCWs (Kang et al., 2020). Online-push messages of mental health self-help and self-help books are mostly preferred by women HCWs (Kang et al., 2020). Measures to support HCWs financially (Almaghrabi et al, 2020), provision of rest areas for sleep and recovery (Yin, Kang et al, 2020), care for basic physical needs such as food (Kang et al, 2020,), training programs to improve resiliency (Zhu et al, 2020), information on protective measures (Kang et al., 2020), and access to leisure activities (Kang et al, 2020) and counsellors (Kang et al, 2020, Liu et al, 2020) are considered potential strategies to support HCWs during a pandemic. However, these studies did not measure the impact of these interventions.

## DISCUSSION

In this rapid scoping review, we summarized evidence to examine common triggers for stress, burnout, and depression in women HCWs and the interventions that can prevent them.

Preliminary findings of causes of increased stress and mental health issues provide and possible strategies for health care organizations to address modifiable factors, such as ongoing training to increase confidence in caring for COVID 19 patients, providing clear infection control guidelines and sufficient PPE, and optimizing working conditions for health care workers.

There was a significant variability in the tools used to measure mental health. This limits the generalizability of our findings. The current literature showed that women HCWs present high levels of anxiety, depression, and burnout. We identified a broad number of common triggers, including individual-level factors, factors relating to work conditions, and systems-level factors. The current literature lacks data on women’s socio-economic, cultural and ethno-racial differences in mental health and resilience.

Further, only a few articles have described interventions to prevent stress, burnout, and depression in women HCWs during pandemics. There is limited evidence regarding the impact of these interventions to reduce stress, burnout, and depression. Mental health services such as online resources, psychological assistance hotlines, and group activities for stress reduction are poorly utilized whereas self-help resources such as books and push messages are mostly preferred by women HCWs. Lastly, there exists a lack of evidence on institutional and systemic practices that facilitate resilience and address HCW mental health.

The current literature has emerged from limited geographical regions. It is not clear how variations in health care and organizational and cultural contexts will shape the outcomes of similar studies spread across a broader geographic area. We expect to see an increased number of publications on the impact of COVID-19 on health professionals over the next several months.

We have registered a rapid review protocol in the International prospective register of systematic reviews (PROSPERO CRD42020189750). As a next step, we will do a subgroup analysis to explore the impact of COVID-19 on mental health issues by professional groups (i.e. doctors, nurses and allied health professionals).

## CONCLUSIONS

We noticed that there is a significant gap in the evidence base. We recommend that health systems decision makers, hospitals, and professional organizations support research studies that measure the long-term impact of COVID-19 on women in health care and support high-quality studies that measure the impact of various mental health interventions and resources supporting women in health care. Given the complex nature of these interventions, we urge future research studies to provide contexts in which the interventions were implemented and mechanisms that shape successful interventions.

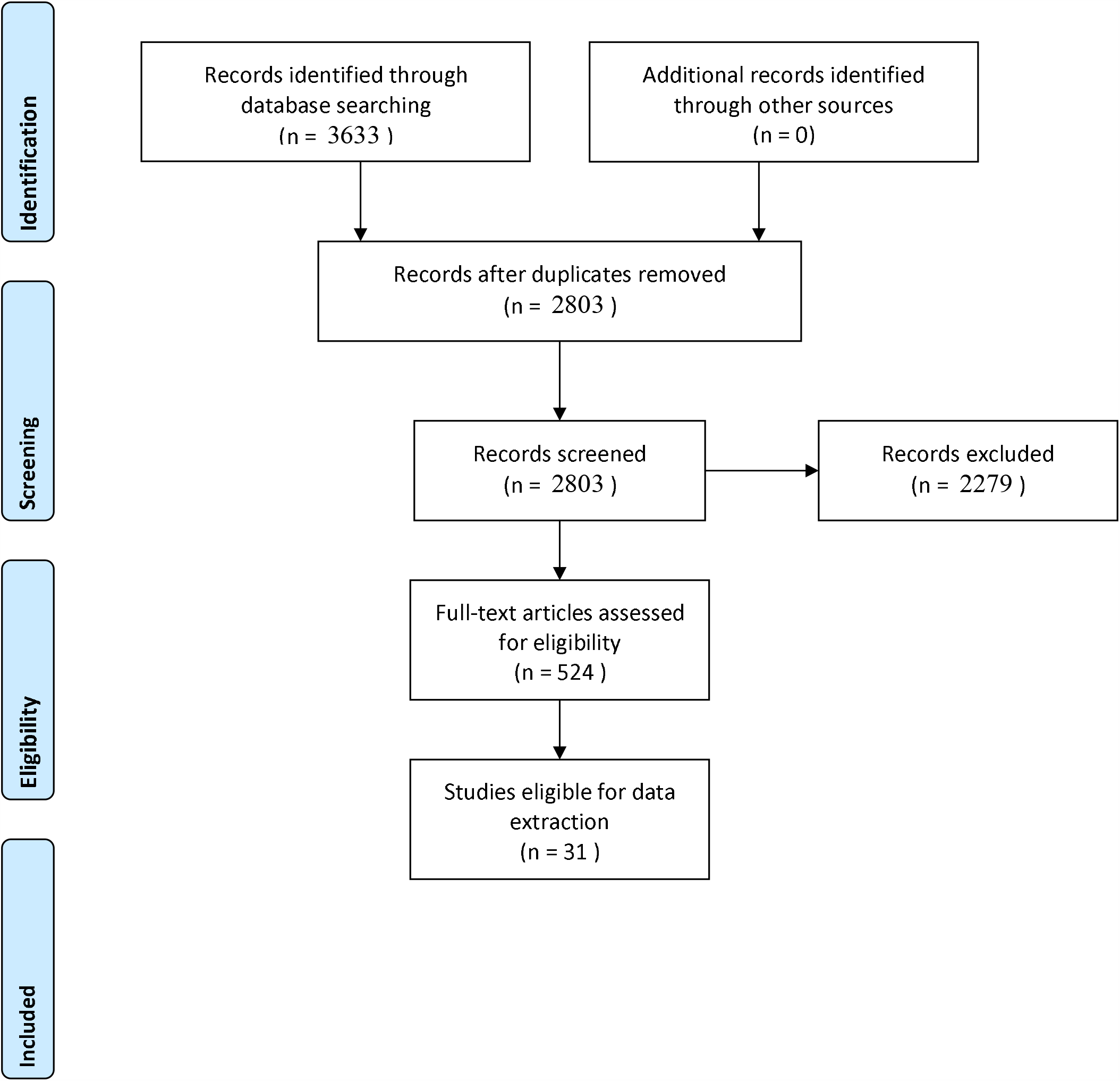
PRISMA 2009 Flow Diagram.

## Data Availability

This review is registered with the Open Science Framework

https://osf.io/y8fdh/?view_only=1d943ec3ddbd4f5c8f6a9290eca2ece7

## Acknowledgement

Authors acknowledge the contribution by Sabine Caleja who helped with article retrieval and screening.

